# Reductions to health-related quality of life associated with cigarette use, e-cigarette use, and depression among US adults

**DOI:** 10.64898/2026.03.19.26348841

**Authors:** Chia-Ling Cheng, Sarah Skolnick, Jamie Tam

## Abstract

**Introduction:** Prior studies suggest e-cigarette use is associated with worse health outcomes, but it remains unclear whether these associations persist independent of cigarette smoking and diagnosed depression and how they jointly affect health-related quality of life (HRQoL). This study provides the first health utility estimates of the independent and combined effects of cigarette use, e-cigarette use, and depression.

**Methods:** We analyzed 2022-2023 Behavioral Risk Factor Surveillance System data, evaluating self-reported physically or mentally unhealthy days. Average unhealthy days were estimated by age, gender, smoking status, depression status, and current e-cigarette use status. Overall healthy days were converted EQ-5D utility scores.

**Results:** Cigarette use, e-cigarette use, and depression were each associated with more mentally and physically unhealthy days, with a more pronounced association observed for mental health. E-cigarette use alone was associated with 2.9-4.2 additional mentally unhealthy days and 1.3-2.8 additional physically unhealthy days per month across adult age groups, with the highest number of additional unhealthy days observed when all three conditions co-occurred. Whether with or without co-diagnosed depression, e-cigarette use was independently associated with poorer mental health among adults aged 18–64. Among young adults, e-cigarette use demonstrated measurable health disutility after accounting for cigarette use and depression.

**Conclusion:** E-cigarette use is independently associated with poorer HRQoL, particularly among younger adults, and these negative associations scale upward when combined with cigarette smoking and depression. Quantifying these joint impacts as health disutility underscores the necessity of integrating e-cigarette cessation into comprehensive tobacco control and mental health policies.

**IMPLICATION:** This study provides the first health utility estimates capturing the independent and combined burdens of cigarette use, e-cigarette use, and depression. By translating population-level behavioral data into EQ-5D utility scores, this study demonstrates that e-cigarette use is independently associated with a measurable reduction in health-related quality of life. These findings provide vital parameters for economic evaluations, highlighting the need to integrate e-cigarette cessation initiatives into broader tobacco control and mental health policies.

## INTRODUCTION

Combustible cigarette smoking is a well-established contributor to both mental and physical health harms.^1–3^ Those who smoke are 1.9 times more likely to experience a major depressive episode in their lifetime.^4^ Furthermore, smoking and depression have a bidirectional relationship, meaning not only does smoking contribute to depression risk, but depression increases the likelihood of taking up and continuing smoking.^5,6^ Smoking is also associated with a host of physical ailments, including cardiovascular disease, chronic obstructive pulmonary disease, and lung cancer, which is the leading cause of cancer deaths.^7,8^ These physical ailments increase the risk of mental health issues, and in turn, mental health issues fuel disease progression.^9,10^ Ultimately, individuals who smoke may be caught in a feedback loop of ill-health and worsening quality of life; smoking cigarettes leads to depression and physical ailments, which in turn lead to further depressive episodes and continued smoking.

While these associations are supported by decades of evidence, the health effects of e-cigarettes remain less well understood. Emerging research has raised concerns about the relationship between e-cigarette use and depressive symptoms, especially among young people, where e-cigarette use is highly prevalent.^9^ Recent studies have observed that those who use e-cigarettes are more likely to report depression or other psychological issues. A study of 2016-2017 Behavioral Risk Factor Surveillance System (BRFSS) data showed a cross-sectional link between e-cigarette use and depression. After controlling for demographic characteristics, heavy alcohol use, and smoking, current e-cigarette users had over twice the odds of having a clinical depression diagnosis compared to never-users.^11^ Other analyses of the U.S. Youth Risk Behavior Survey and a study in Korea have also found higher levels of depression among current e-cigarette users compared to non-users in teens.^12–15^

Given that the prevalence of depression among adolescents increased from 8.2% in 2013 to 13.1% in 2023^16^, and that e-cigarettes have remained the most commonly used tobacco product among youth during this period^17,18^ it is critical to quantify the impact of e-cigarette use on physical and mental health. Furthermore, little is known about the combined effects of cigarette use, e-cigarette use, and depression on overall health status—for example, whether the association between tobacco use and mental health persists even without the presence of depression, and how tobacco use and depression jointly affect physical health outcomes. Therefore, this study aims to examine the interrelationships among these three factors and to quantify their independent and joint effects on population-level health utility scores, with particular focus on e-cigarette use among the younger population.

To quantify these associations in a way that captures their combined impact on overall health and is relevant for policy, we use health utility scores, a widely used measure of health-related quality of life derived from instruments such as the EQ-5D, a standardized questionnaire that captures mobility, self-care, main activities, pain/discomfort, and anxiety/depression.^19,20^ The utility scores form the basis of quality-adjusted life years (QALYs), which are used to evaluate the value and cost-effectiveness of public health interventions. We first used nationally representative BRFSS data to examine physically and mentally healthy days associated with depression, cigarette use, and e-cigarette use. Then we convert these healthy days into a utility score, providing a standardized measure of the impact of these exposures on quality of life.

## METHODS

### Study Participants

To estimate the health impacts of depression, cigarette use, and e-cigarette use, we analyzed 2022 and 2023 Behavioral Risk Factor Surveillance System (BRFSS) data.^21^ The BRFSS is a state-based, nationally representative survey that collects data about health-related risk behaviors, chronic health conditions, and use of preventive services.^21^ The final sample size is 768,707 after excluding missing values for age (N = 16,858), depression status (N = 5,399), cigarette use status (N = 58,524), e-cigarette use status (N = 58,911), physically healthy days (N = 21,712), and mentally healthy days (N = 17,175).

### Depression, cigarette use, and e-cigarette use categories

Depression status was self-reported and measured with the question: “Has a doctor, nurse, or other health professional ever told you that you had a depressive disorder (including depression, major depression, dysthymia, or minor depression)?” Cigarette use status was also self-reported and classified into three categories: current smoker, former smoker, and never smoker. The classification was based on responses to two questions: “Have you smoked at least 100 cigarettes in your entire life?” and “Do you now smoke cigarettes every day, some days, or not at all?” For stratified analyses and graphical presentation, former and never smokers were combined into a single non-current smoker category.

E-cigarette use status was self-reported and categorized into two groups: current users and non-current users. This was assessed with the question: “Would you say you have never used e-cigarettes or other electronic e-cigarette products in your entire life or now use them every day, use them some days, or used them in the past but do not currently use them at all?”

### Main measures

The primary outcomes of this study are self-reported physically healthy days and mentally healthy days, which represent health-related quality of life (HRQoL) as measured by the CDC’s Healthy Days Core Module (HRQoL-4) in the BRFSS.^22^ These outcomes are measured with the following questions: “Now thinking about your physical health, which includes physical illness and injury, for how many days during the past 30 days was your physical health not good?”, and “Now thinking about your mental health, which includes stress, depression and problems with emotions, for how many days during the past 30 days was your physical health not good?”^23^ We calculated the average number of physically and mentally healthy days among adults aged 18 and older, stratified by age, gender, depression status, cigarette use status, and e-cigarette use status, using individuals who use neither cigarette nor e-cigarette and do not have diagnosed depression as the reference group. All analyses accounted for the BRFSS complex sampling design using the provided survey weights to ensure nationally representative estimates.

We then converted the number of overall healthy days into a health utility score ranging from 0 to 1, using the conversion method developed by Jia and Lubetkin.^23^ Primary utility estimates, stratified by current versus non-current smoking status, are presented in Table 2. For completeness, utility scores further stratified by never, former, and current smoking status are provided in the supplemental materials. (Supplement Table 1&2)

All analyses were conducted in R 4.4.2^24^

## RESULT

Table 1 summarizes the demographic characteristics of the study sample from the combined 2022 and 2023 BRFSS survey data, stratified by tobacco use status (cigarette-only, e-cigarette-only, dual use, and neither). There were 768,707 adult participants included in this study, of which 9.9% exclusively smoke cigarettes, 3.8% exclusively use e-cigarettes, 1.6% dual use both products, and the remaining 84.7% who use neither (Table 1). People who used e-cigarettes only were disproportionately younger compared with people who used cigarettes only and people who used both cigarettes and e-cigarettes. People who used both cigarettes and e-cigarettes reported the highest prevalence of depression (43.6%), compared with those who used cigarettes only (30.0%) or e-cigarettes only (37.2%). Overall, people who dual used both cigarettes and e-cigarettes were more likely to be unemployed and reported a greater number of mentally unhealthy days than other tobacco use groups, whereas people who exclusively smoked cigarettes reported the highest number of physically unhealthy days (mean, 6.46 days).

**Table 1.** Demographic characteristics in different cigarette use groups in 2022-2023 BRFSS data. No. (%)

| Characteristic/<br>tobacco use | **Cigarette Only**<br>**(N=75933)** | **E-cigarette**<br>**Only**<br>**(N= 29250)** | **Dual Use**<br>**(N= 12424)** | **Neither**<br>**(N=651100)** |
| --- | --- | --- | --- | --- |
| Age |  |  |  |  |
| **18-24** | 1238 (3.3) | 7519 (34.9) | 1553 (18.9) | 37099 (11.9) |
| **25-44** | 19719 (36.7) | 13650 (47.5) | 5902 (53.2) | 146502 (31.7) |
| **45-64** | 33649 (41.2) | 5827 (14.1) | 3864 (23.5) | 205891 (30.9) |
| **65+** | 21327 (18.8) | 2254 (3.5) | 1105 (4.5) | 261608 (25.5) |
| Gender |  |  |  |  |
| **women** | 37723 (45.5) | 13343 (45.0) | 5808 (43.8) | 348902 (52.5) |
| **men** | 38210 (54.5) | 15907 (55.0) | 6616 (56.2) | 302198 (47.5) |
| Depression<br>status |  |  |  |  |
| **With**<br>**depression** | 23176 (30.0) | 11150 (37.2) | 5443 (43.6) | 120723 (18.2) |
| **Without**<br>**depression** | 52757 (70.0) | 18100 (62.8) | 6981 (56.4) | 530377 (81.8) |
| Race/ethnicity |  |  |  |  |
| **White** | 54357 (62.1) | 20550 (61.4) | 8626 (60.0) | 480774 (58.4) |
| **Black** | 6710 (13.3) | 1749 (9.5) | 790 (9.5) | 47837 (11.5) |
| **Hispanic** | 6218 (14.9) | 3262 (16.5) | 1293 (17.7) | 63256 (19.2) |
| **Other** | 6898 (9.7) | 3147 (12.6) | 1448 (12.8) | 46079 (10.8) |
| Education level |  |  |  |  |
| **Less than**<br>**High School** | 8586 (20.3) | 1573 (8.8) | 1428 (18.8) | 29873 (9.7) |
| **High school**<br>**graduate or**<br>**some college** | 51815 (66.7) | 19437 (72.1) | 8761 (70.0) | 311587 (55.2) |
| **College**<br>**graduate** | 15355 (13.1) | 8189 (19.1) | 2213 (11.2) | 307913 (35.1) |
| Employment<br>status |  |  |  |  |
| **Employed** | 36994 (63.5) | 20545 (75.6) | 7660 (73.3) | 329997 (60.4) |
| **Unemployed** | 7963 (14.7) | 2694 (10.9) | 1629 (16.7) | 47423 (10.4) |
| **Student** | 519 (1.2) | 1818 (9.5) | 353 (4.4) | 16581 (5.4) |
| **Retired** | 18715 (20.7) | 2190 (4.0) | 1081 (5.6) | 222950 (23.7) |
| Marital status |  |  |  |  |
| **Married** | 27667 (38.7) | 9325 (29.7) | 3497 (28.8) | 358590 (53.4) |
| **Divorced** | 17318 (18.5) | 3429 (8.4) | 2328 (14.5) | 75240 (9.1) |
| **Widowed** | 8665 (8.6) | 820 (1.5) | 656 (3.4) | 71274 (7.1) |
| **Single** | 21856 (34.3) | 15492 (60.4) | 5852 (53.2) | 142281 (30.4) |
| **Physically**<br>**unhealthy**<br>**days** | 6.46 (10.44) | 4.34 (8.18) | 6.31 (9.96) | 3.81 (8.05) |
| **Mentally**<br>**Unhealthy**<br>**days** | 7.36 (10.74) | 9.05 (10.65) | 10.75 (11.76) | 4.23 (8.00) |

### Mentally Unhealthy Days

Across all age groups, those who do not use cigarettes or e-cigarettes and do not have depression have the least number of mentally unhealthy days, and those who smoke, vape, and have depression have the greatest number of mentally unhealthy days. E-cigarette use alone was associated with 2.9-4.2 (95% CI: 2.0-4.6) additional mentally unhealthy days per month across all age groups, independent of cigarette use and depression status. There were no major differences by gender overall. However, the mental health burden associated with cigarette use, e-cigarette use, and depression combined was substantially higher among women aged 18-24 compared with men in the same group, and higher among men aged 65 and older compared with women of the same age.

Depression status is self-reported by individuals who have been told they have a depression diagnosis, whereas mentally unhealthy days may capture both undiagnosed and diagnosed depression. As expected, we observed a strong relationship between diagnosed depression and mentally unhealthy days. However, even in the absence of diagnosed depression, tobacco use was still associated with worse mental health. Among individuals aged 18-64 without depression, those who did not use cigarettes or e-cigarettes reported the fewest mentally unhealthy days. Compared with this reference group, people who used e-cigarettes only reported 1.9-2.2 (95% CI: 1.2-3.1) additional mentally unhealthy days among men and 2.3-2.8 (95% CI: 1.5-3.7) among women (Figure 1). Additionally, compared with the same reference group, men and women who used both e-cigarettes and cigarettes reported an additional 2.4-3.6 (95% CI: 1.4-4.9) and 3.5-5.1 (95% CI: 2.2-8.4) extra mentally unhealthy days, respectively. (Figure 1) In the presence of diagnosed depression, on the other hand, e-cigarette use remained associated with an additional mental health burden. Among individuals aged 18-64 with depression, men and women who use e-cigarettes only reported an additional 2.1-2.7 (95% CI: 0.5-4.0) and 2.0-3.8 (95% CI: 1.0-5.0) mentally unhealthy days compared to those who use neither tobacco product. When all three conditions were present, the greatest mental health burden was observed, with men and women reporting 10.5-19.3 (95% CI: 8.2-25.6) and 9.8-15.2 (95% CI: 7.2-17.3) additional mentally unhealthy days, respectively, compared with individuals with none of these conditions.

**Figure 1.**
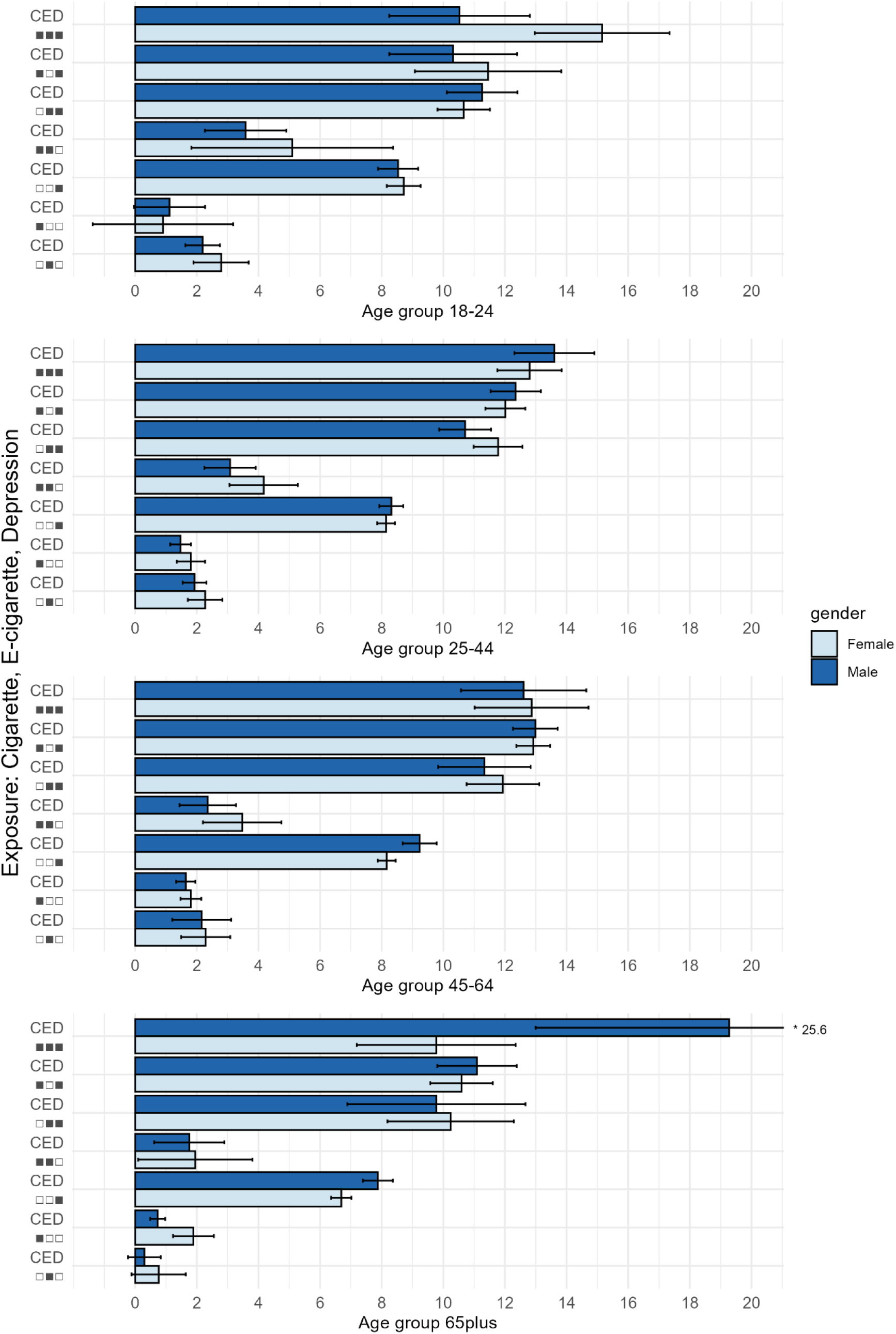
Mean Number of Additional Mentally Unhealthy Days by Cigarette use, E-cigarette use, and Depression Status

### Physically Unhealthy Days

Overall, physically unhealthy days increased with the number of co-occurring conditions, although this pattern was less consistent than that observed for mentally unhealthy days. Nevertheless, associations between e-cigarette use and physically unhealthy days followed a similar pattern, albeit with smaller magnitudes. Across all age groups, e-cigarette use was associated with 1.3-2.8 (95% CI: 1.0-3.8) additional physically unhealthy days per month, independent of cigarette use and depression status. The burden of physically unhealthy days increased when e-cigarette use co-occurred with cigarette use or depressive symptoms.

In particular, among adults aged 18-24, men and women who used both cigarettes and e-cigarettes without a history of diagnosed depression reported an additional 1.4 (95% CI: 0.5-2.3) and 3.9 (95% CI: 1.0-6.8) physically unhealthy days, respectively, compared with individuals with none of these conditions. Among those who reported diagnosed depression and used e-cigarettes only, men and women reported 2.9 (95% CI: 2.0-3.8) and 3.1 (95% CI: 2.5-3.6) additional physically unhealthy days, respectively. When all three conditions were present, the greatest physical health burden was observed among adults aged 18-24, with men and women reporting 4.9 (95% CI: 3.1-6.6) and 5.9 (95% CI: 3.9-8.0) additional physically unhealthy days, respectively, compared with individuals with none of these conditions. (Figure 2)

**Figure 2.**
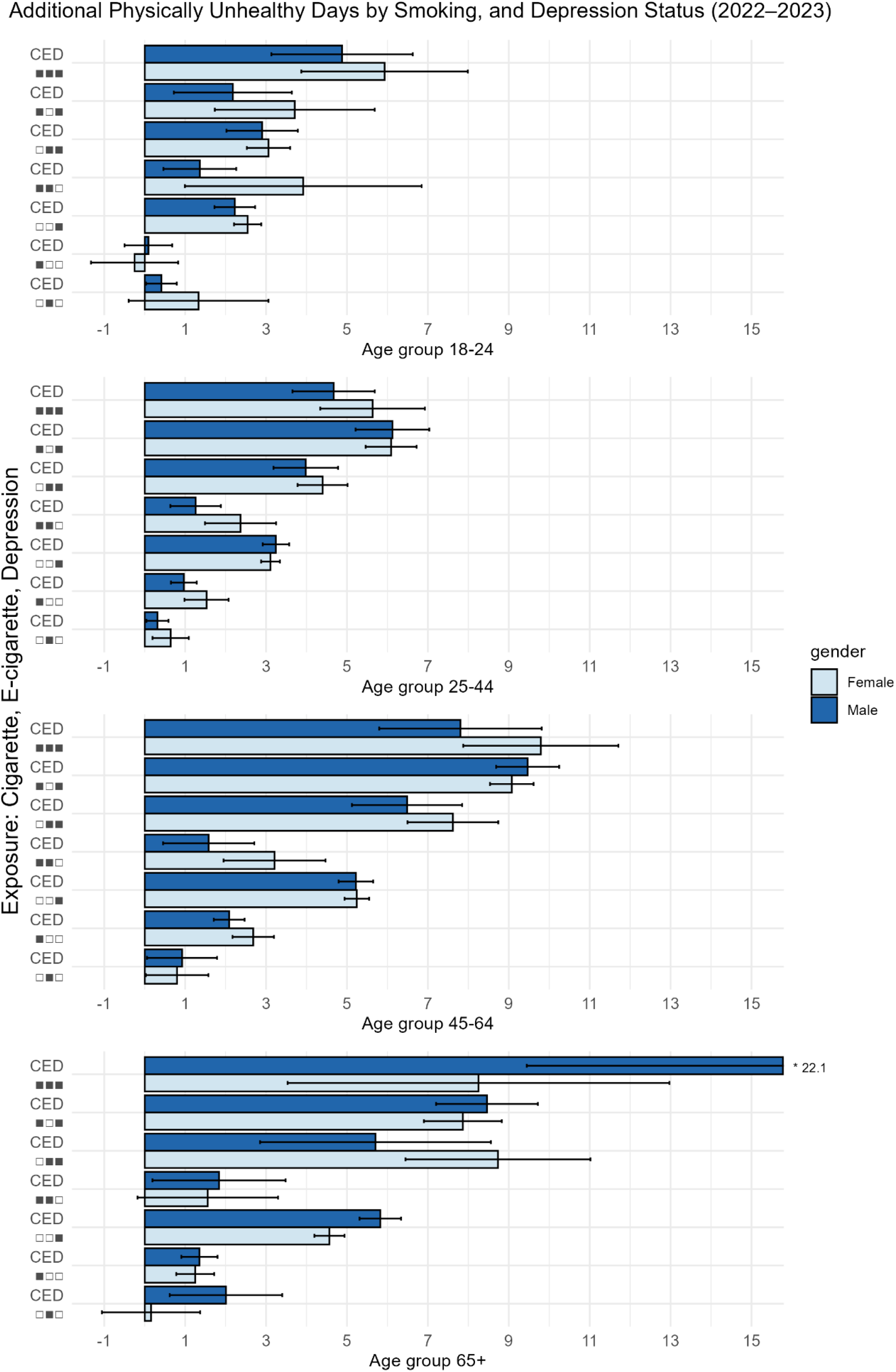
Mean Number of Additional Physically Unhealthy Days by Cigarette use, E-cigarette use, and Depression Status

There were no major differences by gender overall. However, the physical health burden associated with cigarette use, e-cigarette use, and depression together was substantially higher among women aged 18-24 compared with men in the same group, and higher among men aged 65 and older compared with women of the same age.

### Combined Physically and Mentally Unhealthy Days

The association between e-cigarette use and combined physically and mentally unhealthy days mirrored the patterns observed for physical and mental unhealthy days separately.Individuals who neither use cigarettes nor e-cigarettes nor have a history of diagnosed depression reported the highest utility scores. E-cigarette use alone was associated with modest utility losses. Among adults aged 18-24 without cigarette use or diagnosed depression, e-cigarette use was associated with disutility scores of 0.011 in men and 0.015 in women. In the same age group, e-cigarette use combined with cigarette use was associated with disutility scores of 0.014 in men and 0.022 in women, whereas e-cigarette use combined with diagnosed depression was associated with disutility scores of 0.044 in men and 0.042 in women. When all three conditions were present, disutility scores increased to 0.044 in men and 0.06 in women. (Table 2) Similar associations were observed in the rest of the age groups, with e-cigarette use linked to small but consistent disutility scores among men and women who neither use cigarettes nor have a history of diagnosed depression, or who used e-cigarettes in combination with one of these conditions. However, the associations between e-cigarette use and combined physically and mentally unhealthy days were less clear when both smoking and depression were present across all age groups. Utility scores stratified by never, former, and current smoking status are provided in the supplement.

**Table 2.**
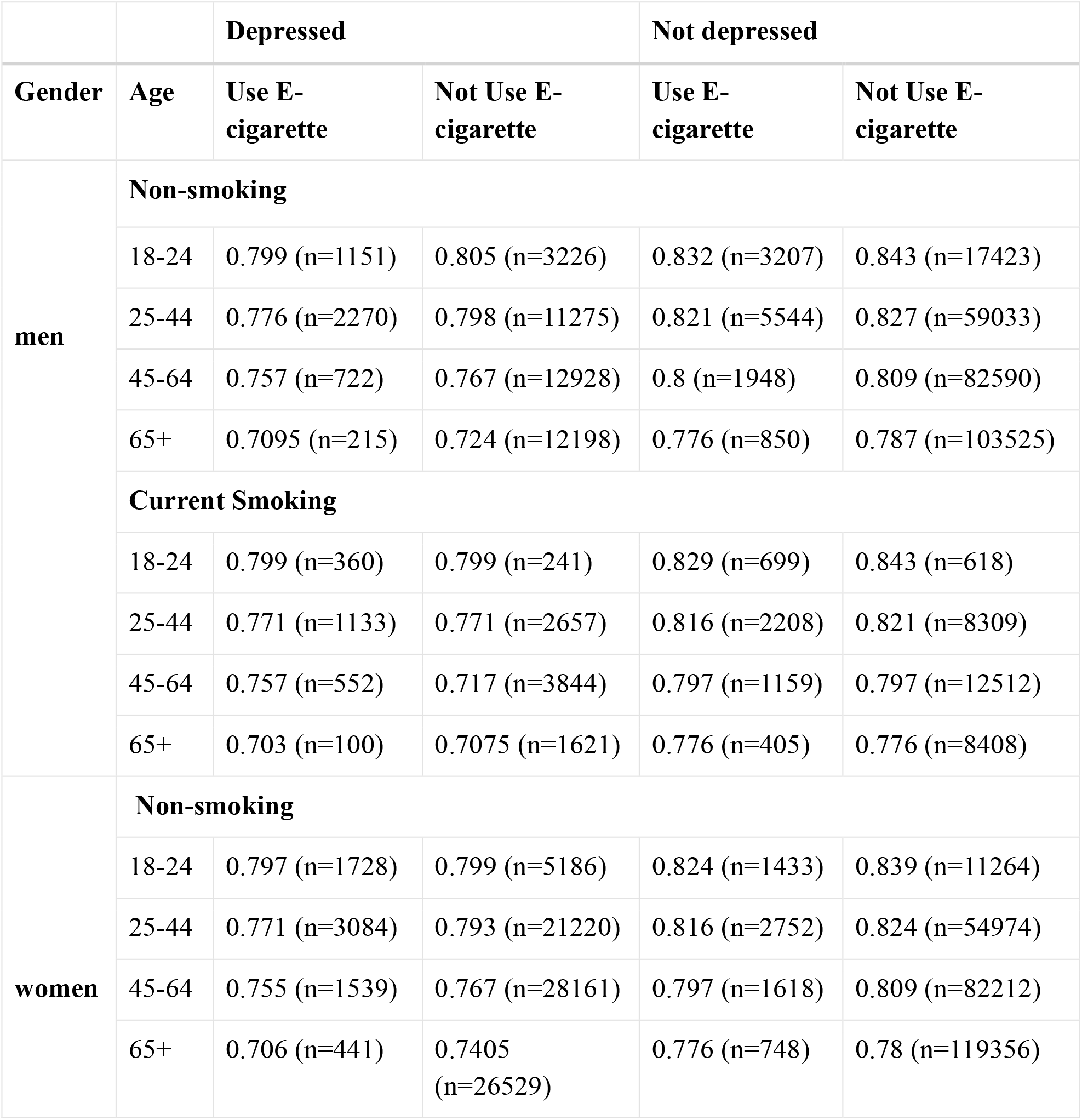
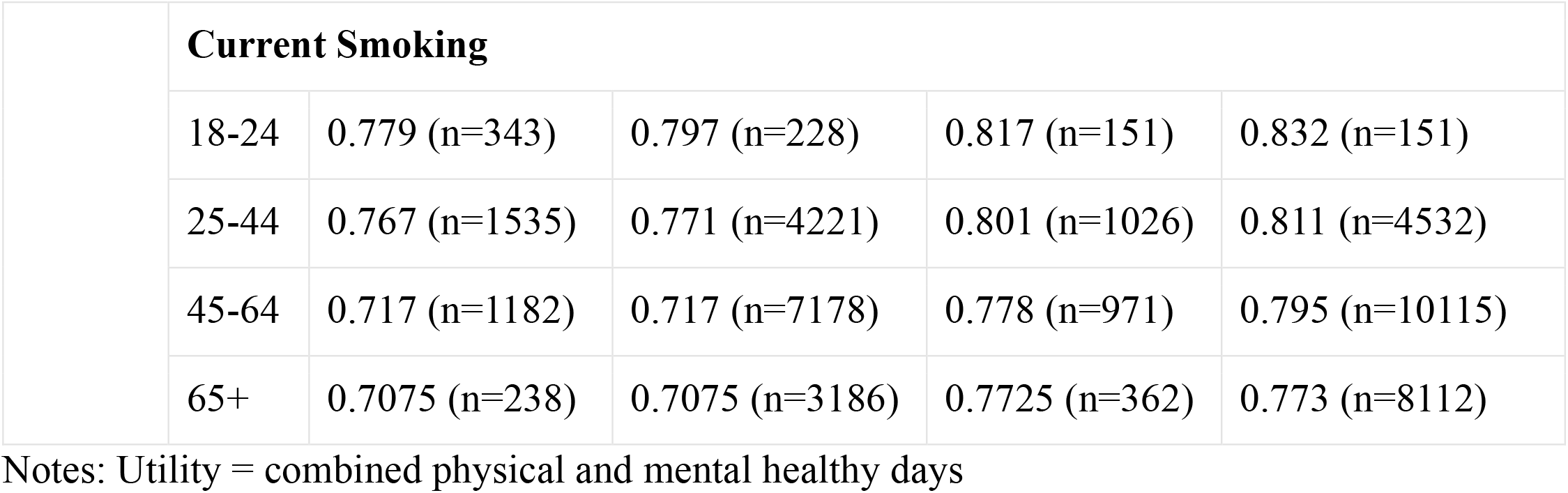
Utility scores by smoking, depression, and e-cigarette status among adults 18+.

## DISCUSSION

This study elucidates the independent and combined associations of e-cigarette use, cigarette use, and depression with physically and mentally unhealthy days. We furthermore quantify the joint effects of all three factors on individual health utility scores. Among individuals without a history of diagnosed depression, e-cigarette use was associated with an increased number of mentally unhealthy days. Among those with diagnosed depression, e-cigarette use was associated with an additional increase in mental health burden beyond the elevated baseline associated with depression in the same age group. This finding aligns with previous research showing that those who use e-cigarettes are more likely to report depression and other psychological symptoms.^11,12^ Given evidence that the depression rate has increased among young adults over the past decade^16,25^, and e-cigarettes remained the most commonly used tobacco product in this population^17,18^, young adults may be particularly vulnerable to the adverse mental health effects associated with e-cigarette use. This is consistent with neurobiological evidence showing that adolescents and young adults are uniquely vulnerable to the effects of nicotine on the developing brain systems involved in reward and mood regulation.^26,27^

While the mental health impacts of e-cigarette use were more pronounced and consistent, the associations with physically unhealthy days were smaller in magnitude and more heterogeneous across age and gender groups. Nevertheless, our findings suggest that e-cigarette use may contribute to modest increases in physically unhealthy days at the population level. Prior studies have shown that e-cigarette use is associated with psychological symptoms such as depressive disorders,^11,12^ which in turn are linked to poorer physical health outcomes.^28–30^ We observed that physically unhealthy days increased substantially when depressive symptoms were present, regardless of tobacco use status. This suggests that depression is a strong determinant of physical health, which may overshadow the more modest contribution of e-cigarette use, making its association with physical health outcomes less consistent and harder to detect, particularly in subgroup analyses.

The effect of e-cigarette use on health has been debated since the introduction of e-cigarettes. While some studies suggest that e-cigarettes may be less harmful than combustible tobacco^31,32^, substantial uncertainty remains regarding their overall health impact, particularly with respect to long-term use and their relationship with mental health outcomes. ^32^ Our findings suggest that when cigarette use, particularly in combination with depression, dominates overall health-related quality of life, rendering the incremental impact of e-cigarette use less distinct.Nevertheless, e-cigarette use was still associated with measurable reductions in health-related quality of life across age groups, with the most pronounced effects observed among younger adults.

Utilities are essential inputs for cost-effectiveness and simulation modeling analysis. This study provides utility-relevant evidence on how e-cigarette use, cigarette use, and depression jointly affect self-reported health. Notably, the growing prevalence of e-cigarette use among young people,^25^ coupled with the lack of established treatment options for vaping cessation,raises important public health concerns. Our findings suggest that e-cigarette use, both independently and in combination with cigarette use or depression, is meaningfully associated with reduced health-related quality of life. Integrating these utility impacts into future economic evaluations can inform more effective resource allocation and guide policies that address both the physical and mental health consequences of e-cigarette use, particularly among young adults. Considering that it may take decades to understand the long-term health risks of vaping, our evaluations of short-term health harms in the form of utility scores can be integrated with broader evaluations of the population health impacts of tobacco control and vaping interventions. Many existing modeling analyses of tobacco control interventions report health outcomes related to mortality and life-years gained^33,34^, meaning that potential reductions in quality of life associated with current vaping are often not fully incorporated into the primary health outcomes evaluated. This analysis offers a first step toward addressing that gap. Future studies of the short and long-term impacts of interventions can rely on the utility estimates reported here to reflect quality of life and more accurately capture the scope of health harms and morbidities linked to both cigarette and e-cigarette use. In separate analyses, the authors have applied these estimates to evaluate the health equity and economic impacts of a nicotine product standard in the US.^35^

### Limitations

Due to the observational nature of BRFSS data, we were unable to establish temporal or causal relationships among cigarette use, e-cigarette use, depression, and the number of physically or mentally healthy days. Second, because most variables were self-reported, the findings may be subject to recall bias. However, previous studies have demonstrated that self-reported measures provide a valid assessment of tobacco use.^36^ Third, people who never smoked and formerly smoked were combined into a single reference group to simplify subgroup comparisons, which may obscure meaningful differences in health outcomes. However, to ensure accurate conversion of health utility scores, we provided the disaggregated estimates for never, current, and former smokers in the supplementary materials. Despite these limitations, the BRFSS remains a nationally representative survey that provides reliable estimates for examining health risk behaviors such as cigarette use, e-cigarette use, and depression and their association with health-related quality of life (HRQoL).^36^ Other national surveys include some of these measures but do not capture this combination as consistently, making BRFSS particularly well-suited for this analysis.

In summary, this study provides the first health utility estimates of the independent and combined effects of cigarette use, e-cigarette use, and depression on HRQoL, as measured by physically and mentally unhealthy days. While smoking and depression exhibited the strongest negative effects on both physical and mental well-being, e-cigarette use alone was also associated with worse health outcomes and notable health disutility, particularly among younger adults. These findings reinforce the established understanding of smoking and depression as major public health burdens, while also highlighting emerging concerns associated with both e-cigarette use and depressive symptoms in youth populations. Future efforts should prioritize interventions specifically aimed at the young population, who are disproportionately vulnerable to the health risks associated with e-cigarette use.

## Data Availability

All data used in this study are publicly available from the Behavioral Risk Factor Surveillance System (BRFSS) through the U.S. Centers for Disease Control and Prevention (CDC) at https://www.cdc.gov/brfss/

## Acknowledgments

This work was originally initiated while Ms. Cheng was an MPH student at the Department of Health Policy and Management at the Yale School of Public Health from 2023 to 2025.

## Ethical Statement

This study used publicly available, de-identified data from the Behavioral Risk Factor Surveillance System (BRFSS) and did not involve human subjects as defined by federal regulations.

## Data Availability

The data and analysis code underlying this article will be made publicly available in a GitHub repository upon formal publication.

## Reference

1. Health and Economic Benefits of Tobacco Use Interventions | National Center for Chronic Disease Prevention and Health Promotion (NCCDPHP) | CDC. Accessed June 28, 2025. https://www.cdc.gov/nccdphp/priorities/tobacco-use.html

2. Wu Z, Yue Q, Zhao Z, et al. A cross-sectional study of smoking and depression among US adults: NHANES (2005–2018). Front Public Health. 2023;11. doi:10.3389/FPUBH.2023.1081706,

3. Becker TD, Arnold MK, Ro V, Martin L, Rice TR. Systematic Review of Electronic Cigarette Use (Vaping) and Mental Health Comorbidity among Adolescents and Young Adults. Nicotine and Tobacco Research. 2021;23(3):415–425. doi:10.1093/NTR/NTAA171,

4. Breslau N, Peterson EL, Schultz LR, Chilcoat HD, Andreski P. Major Depression and Stages of Smoking: A Longitudinal Investigation. Arch Gen Psychiatry. 1998;55(2):161–166.doi:10.1001/ARCHPSYC.55.2.161

5. Lechner W V., Janssen T, Kahler CW, Audrain-McGovern J, Leventhal AM. Bi-directional associations of electronic and combustible cigarette use onset patterns with depressive symptoms in adolescents. Prev Med (Baltim). 2016;96:73. doi:10.1016/J.YPMED.2016.12.034

6. Chaiton MO, Cohen JE, O’Loughlin J, Rehm J. A systematic review of longitudinal studies on the association between depression and smoking in adolescents. BMC Public Health. 2009;9(1):1–11. doi:10.1186/1471-2458-9-356/TABLES/3

7. Services USD of H and H, Prevention C for DC and, Promotion NC for CDP and H, Health O on S and. The Health Consequences of Smoking—50 Years of Progress. Published online 2014:1–36. doi:NBK179276

8. Siegel RL, Giaquinto AN, Jemal A. Cancer statistics, 2024. CA Cancer J Clin. 2024;74(1):12–49. doi:10.3322/CAAC.21820

9. Dragioti E, Radua J, Solmi M, et al. Impact of mental disorders on clinical outcomes of physical diseases: an umbrella review assessing population attributable fraction and generalized impact fraction. World Psychiatry. 2023;22(1):86. doi:10.1002/WPS.21068

10. Momen NC, Østergaard SD, Heide-Jorgensen U, Sørensen HT, McGrath JJ, Plana-Ripoll O. Associations between physical diseases and subsequent mental disorders: a longitudinal study in a population-based cohort. World Psychiatry. 2024;23(3):421. doi:10.1002/WPS.21242

11. Obisesan OH, Mirbolouk M, Osei AD, et al. Association Between e-Cigarette Use and Depression in the Behavioral Risk Factor Surveillance System, 2016-2017. JAMA Netw Open. 2019;2(12):e1916800. doi:10.1001/JAMANETWORKOPEN.2019.16800

12. Javed S, Usmani S, Sarfraz Z, et al. A Scoping Review of Vaping, E-Cigarettes and Mental Health Impact: Depression and Suicidality. J Community Hosp Intern Med Perspect. 2022;12(3):33. doi:10.55729/2000-9666.1053

13. Comparison of Emotional and Psychological Indicators According to the Presence or Absence of the Use of Electronic Cigarettes among Korea Youth Smokers − ProQuest. Accessed January 1, 2026. https://www.proquest.com/openview/046155cf9ade14a7312bc122fcf0f466/1?pq-origsite=gscholar&cbl=936334

14. Chadi N, Li G, Cerda N, Weitzman ER. Depressive Symptoms and Suicidality in Adolescents Using e-Cigarettes and Marijuana: A Secondary Data Analysis From the Youth Risk Behavior Survey. JAddict Med. 2019;13(5):362–365. doi:10.1097/ADM.0000000000000506

15. Lee Y, Lee KS. Association of Depression and Suicidality with Electronic and Conventional Cigarette Use in South Korean Adolescents. Subst Use Misuse. 2019;54(6):934–943.doi:10.1080/10826084.2018.1552301

16. Brody DJ, Hughes JP. Key findings Data from the National Health and Nutrition Examination Survey. Published online 2021. Accessed November 6, 2025.https://www.cdc.gov/nchs/products/index.htm.

17. Bandi P, Cahn Z, Goding Sauer A, et al. Trends in E-Cigarette Use by Age Group and Combustible Cigarette Smoking Histories, U.S. Adults, 2014–2018. Am J Prev Med. 2021;60(2):151–158.doi:10.1016/j.amepre.2020.07.026

18. Osterman AV, Briones E, Jamal A, Marynak K, Valenzuela C. Electronic Cigarette Use Among Adults in the United States. Published online January 30, 2025. doi:10.15620/cdc/174583

19. Fanshel S, Bush JW. EuroQol - a new facility for the measurement of health-related quality of life. Health Policy (New York). 1990;16(3):199–208. doi:10.1016/0168-8510(90)90421-9

20. Herdman M, Gudex C, Lloyd A, et al. Development and preliminary testing of the new five-level version of EQ-5D (EQ-5D-5L). Qual Life Res. 2011;20(10):1727–1736. doi:10.1007/s11136-011-9903-x

21. U.S. Department of Health and Human Services. Centers for Disease Control and Prevention Behavioral risk factor surveillance system (BRFSS) data 2022. 2022. Accessed April 23, 2025. https://www.cdc.gov/brfss/annual_data/annual_2022.html

22. for Disease Control C. Measuring Healthy Days Population Assessment of Health-Related Quality of Life. Published online 2000. Accessed July 17, 2025. http://www.cdc.gov/nccdphp/brfss/

23. Jia H, Lubetkin EI. Estimating EuroQol EQ-5D scores from population healthy days data. Medical Decision Making. 2008;28(4):491–499. doi:10.1177/0272989X07312708,

24. R: The R Project for Statistical Computing. Accessed April 29, 2026. https://www.r-project.org/

25. Moustafa AF, Testa S, Rodriguez D, Pianin S, Audrain-McGovern J. Adolescent depression symptoms and e-cigarette progression. Drug Alcohol Depend. 2021;228:109072.doi:10.1016/J.DRUGALCDEP.2021.109072

26. Yuan M, Cross SJ, Loughlin SE, Leslie FM. Nicotine and the adolescent brain. J Physiol. 2015;593(Pt 16):3397. doi:10.1113/JP270492

27. John U, Meyer C, Rumpf HJ, Hapke U. Smoking, nicotine dependence and psychiatric comorbidity-A population-based study including smoking cessation after three years. Drug Alcohol Depend. 2004;76(3):287–295. doi:10.1016/j.drugalcdep.2004.06.004

28. Mezuk B, Eaton WW, Albrecht S, Golden SH. Depression and Type 2 Diabetes Over the LifespanA meta-analysis. Diabetes Care. 2008;31(12):2383–2390. doi:10.2337/DC08-0985

29. Pan A, Keum N, Okereke OI, et al. Bidirectional Association Between Depression and Metabolic SyndromeA systematic review and meta-analysis of epidemiological studies. Diabetes Care. 2012;35(5):1171–1180. doi:10.2337/DC11-2055

30. Chen X, Liu X, Li F, et al. Depression and health outcomes: An umbrella review of systematic reviews and meta-analyses of observational studies. Translational Psychiatry 2025 15:1. 2025;15(1):1–17. doi:10.1038/s41398-025-03463-8

31. Nutt DJ, Phillips LD, Balfour D, et al. Estimating the harms of nicotine-containing products using the MCDA approach. Eur Addict Res. 2014;20(5):218–225. doi:10.1159/000360220,

32. E-cigarettes around 95% less harmful than tobacco estimates landmark review - GOV.UK. Accessed July 27, 2025. https://www.gov.uk/government/news/e-cigarettes-around-95-less-harmful-than-tobacco-estimates-landmark-review

33. Holford TR, Meza R, Warner KE, et al. Tobacco Control and the Reduction in Smoking-Related Premature Deaths in the United States, 1964-2012. JAMA. 2014;311(2):164–171.doi:10.1001/jama.2013.285112

34. Levy DT, Mabry PL, Graham AL, Orleans CT, Abrams DB. Exploring Scenarios to Dramatically Reduce Smoking Prevalence: A Simulation Model of the Three-Part Cessation Process. Am J Public Health. 2010;100(7):1253. doi:10.2105/AJPH.2009.166785

35. Skolnick S, Brouwer A, Cheng C ling, Tam J. Health, Equity, and Economic Impacts of a Nicotine Product Standard in the United States for People With and Without Major Depression. medRxiv. Published online November 5, 2025:2025.07.10.25331302. doi:10.1101/2025.07.10.25331302

36. Hrywna M, Manderski MTB, Delnevo CD. Sex Differences in the Association of Psychological Distress and Tobacco Use. Am J Health Behav. 2014;38(4):570. doi:10.5993/AJHB.38.4.10

