## Supplemental Table 1-2 for "Reductions to health-related quality of life associated with cigarette use, e-cigarette use, and depression among US adults"

Supplement

Table 1. Utility scores by cigarette use, depression, and e-cigarette use status - men ages 18+

| | Depressed | | Not depressed | |
|---|---|---|---|---|
| Age | Use E-cigarette | Not Use E-cigarette | Use E-cigarette | Not Use E-cigarette |
| Never smoking | | | | |
| 18-24 | 0.799 (n=781) | 0.805 (n=2995) | 0.832 (n=2525) | 0.843 (n=16677) |
| 25-44 | 0.781 (n=659) | 0.798 (n=7684) | 0.824 (n=1991) | 0.827 (n=45413) |
| 45-64 | 0.759 (n=135) | 0.773 (n=7589) | 0.795 (n=38) | 0.818 (n=57979) |
| 65+ | 0.724 (n=41) | 0.7405 (n=5540) | 0.792 (n=185) | 0.78 (n=55421) |
| Current smoking | | | | |
| 18-24 | 0.799 (n=360) | 0.799 (n=241) | 0.829 (n=699) | 0.843 (n=618) |
| 25-44 | 0.771 (n=1133) | 0.771 (n=2657) | 0.816 (n=2208) | 0.821 (n=8309) |
| 45-64 | 0.757 (n=552) | 0.717 (n=3844) | 0.797 (n=1159) | 0.797 (n=12512) |
| 65+ | 0.703 (n=100) | 0.7075 (n=1621) | 0.773 (n=405) | 0.776 (n=8408) |
| Former smoking | | | | |
| 18-24 | 0.797 (n=370) | 0.797 (n=231) | 0.829 (n=682) | 0.843 (n=746) |
| 25-44 | 0.776 (n=1611) | 0.793 (n=3591) | 0.821 (n=3553) | 0.827 (n=13620) |
| 45-64 | 0.757 (n=587) | 0.761 (n=5339) | 0.8 (n=1567) | 0.803 (n=24611) |
| 65+ | 0.7095 (n=174) | 0.724 (n=6658) | 0.776 (n=665) | 0.78 (n=48104) |

Notes: Utility = combined physical and mental healthy days

Table 2. Utility scores by cigarette use, depression, and e-cigarette status - women ages 18+

| | Depressed | | Not depressed | |
|---|---|---|---|---|
| Age | Use E-cigarette | Not Use E-cigarette | Use E-cigarette | Not Use E-cigarette |
| Never smoking | | | | |
| 18-24 | 0.797 (n=1324) | 0.799 (n=4927) | 0.826 (n=1214) | 0.839 (n=11063) |
| 25-44 | 0.776 (n=902) | 0.793 (n=16036) | 0.821 (n=1010) | 0.827 (n=47255) |

| | | | | |
|---|---|---|---|---|
| 45-64 | 0.757 (n=199) | 0.767 (n=18166) | 0.797 (n=287) | 0.809 (n=62577) |
| 65+ | 0.7405 (n=67) | 0.706 (n=15620) | 0.792 (n=170) | 0.787 (n=80496) |
| Current smoking | | | | |
| 18-24 | 0.779 (n=343) | 0.797 (n=228) | 0.817 (n=151) | 0.832 (n=151) |
| 25-44 | 0.767 (n=1535) | 0.771 (n=4221) | 0.801 (n=1026) | 0.811 (n=4532) |
| 45-64 | 0.717 (n=1182) | 0.717 (n=7178) | 0.778 (n=971) | 0.795 (n=10115) |
| 65+ | 0.7075 (n=238) | 0.7075 (n=3186) | 0.7725 (n=362) | 0.773 (n=8112) |
| Former smoking | | | | |
| 18-24 | 0.797 (n=404) | 0.799 (n=259) | 0.821 (n=219) | 0.829 (n=201) |
| 25-44 | 0.771 (n=2182) | 0.781 (n=5184) | 0.811 (n=1742) | 0.821 (n=7719) |
| 45-64 | 0.717 (n=1340) | 0.761 (n=9995) | 0.797 (n=1331) | 0.8 (n=19635) |
| 65+ | 0.706 (n=374) | 0.7405 (n=10909) | 0.776 (n=578) | 0.776 (n=38860) |
